# Acceptability of an intervention to improve uptake of evidence-based emergency myocardial infarction care in Tanzania: A qualitative study

**DOI:** 10.64898/2026.04.07.26348549

**Authors:** Spencer F Sumner, Francis M Sakita, Kelvin F Haukila, Lisa Wanda, Godfrey L Kweka, Jerome J Mlangi, Pankrasi Shayo, Tumsifu G Tarimo, Simran Khanna, Claire Wang, Abigail Pyne, Preeti Manavalan, Nathan M Thielman, Janet P Bettger, Julian T Hertz

## Abstract

Acute myocardial infarction (AMI) is an increasing cause of morbidity and mortality in Sub-Saharan Africa (SSA) but is often underdiagnosed and undertreated. To address this gap, the Multicomponent Intervention to Improve Myocardial Infarction Care (MIMIC) was developed and implemented in the emergency department (ED) of a regional referral center in northern Tanzania.

We conducted in-depth interviews with 20 key stakeholders (physicians, nurses, administrators, and patients) who participated in MIMIC during the first year of implementation. Purposive sampling was used to recruit a broad range of participants. Interviews were guided by a semi-structured interview guide informed by the Theoretical Framework of Acceptability (TFA). Interview transcripts were thematically analyzed by a team of coders using an inductive, grounded theory approach guided by the seven TFA domains.

Nineteen major themes emerged across all TFA domains. Overall, participants described MIMIC as highly acceptable, minimally burdensome, and well-aligned with professional and ethical values. Perceived effectiveness was most emphasized, with staff citing improvements in AMI recognition, ECG and troponin testing, and use of evidence-based therapies. All components were highlighted as effective and easily integrated into existing workflows. Patients valued the educational pamphlet for improving knowledge and self-efficacy, though staff expressed concerns about distributing it during acute care, contributing to inconsistent delivery. Champions were viewed as key in promoting adherence and sustaining implementation of the intervention.

MIMIC was widely acceptable in all seven TFA domains among ED providers and patients, with perceived effectiveness driving positive attitudes across stakeholder groups. Use of a co-design approach in MIMIC development likely contributed to high intervention acceptability. Patient education strategies may require adaptation to improve fidelity. These findings suggest that continued implementation and future adaptation of MIMIC may be feasible.

## Introduction

The leading cause of death globally is ischemic heart disease.(1) The incidence of acute myocardial infarction (AMI) has been found to be increasing in Sub-Saharan Africa (SSA),(2) where a growing body of evidence has highlighted opportunities to improve AMI care.(3) In northern Tanzania, for example, recent observational studies found that AMI is frequently misdiagnosed and inadequately treated, due to many factors including limited provider training, ill-equipped facilities, and patient non-adherence to medication.(4,5)These observed gaps in evidence-based AMI care in Tanzania are associated with high mortality, with some studies from the country reporting 30-day mortality of over 40% percent, as compared to the single-digit mortality rates found in high-income countries.(6,7)

In an effort to address these concerns, a multicomponent intervention to improve myocardial infarction care (MIMIC) was implemented at an emergency department in Tanzania.(8,9) MIMIC was developed by an international, interdisciplinary team comprised of implementation scientists, physicians, nurses, patients, and social scientists. This intervention consisted of five main components: (a) provider training on recognizing and treating AMI, (b) triage cards to flag patients who may have AMI, (c) provider pocket cards detailing AMI diagnosis and management, (d) appointed champions to ensure compliance with the intervention, and (e) patient education on AMI via physical and digital pamphlets. Beginning in 2023, the MIMIC intervention was implemented in a pilot trial in the emergency department (ED) at Kilimanjaro Christian Medical Centre (KCMC), in northern Tanzania, to improve uptake of evidence-based AMI care.(9)

Preliminary evidence from this pilot trial found that implementation of MIMIC was associated with significant improvements in AMI testing, AMI case detection, AMI treatment, and uptake of evidence-based secondary preventative therapies.(10) As with all quality improvement interventions, it is important to determine if the intervention was successful not only through efficacy outcomes, but also through acceptability measures. A previous survey of ED staff using the Acceptability of Intervention Measurement (AIM) and Feasibility of Intervention Measurement (FIM) found that the MIMIC intervention was highly acceptable and feasible from the perspective of ED providers.(11,12) A subsequent follow-up study among KCMC ED clinicians found high perceived organizational capacity for sustainability and strong normalization of MIMIC into routine clinical practice, measured using the Clinical Sustainability Assessment Tool (CSAT) and the NoMAD questionnaire.(13) While these quantitative measures provide important insight into implementation outcomes, they do not fully characterize how stakeholders experienced the intervention or which factors shaped acceptability in practice.

Therefore, we used qualitative methods to examine perceptions of MIMIC among ED physicians, nurses, administrators, and patients and to provide contextual insight to inform future adaptation and scale-up. We conducted in-depth interviews with key stakeholders working in, and receiving care from, the KCMC ED during implementation to better understand intervention acceptability.

## Materials and Methods

### Study Design

This study employed a qualitative research design to assess the acceptability and feasibility of the MIMIC intervention in the emergency department (ED) at Kilimanjaro Christian Medical Centre (KCMC).(14) Semi-structured interviews were conducted with key stakeholders over the one-year study period to explore their perceptions, experiences, and attitudes toward the intervention.

### Setting

KCMC is the Zonal Referral Hospital for northern Tanzania, functioning as a major tertiary care and teaching hospital serving a patient population of roughly 11 million people. The MIMIC intervention was implemented in the KCMC ED beginning on September 1^st^, 2023, and interviews with key stakeholders were conducted from January through September 2024. At the time of the interviews, KCMC lacked a cardiologist on staff and did not have capacity for percutaneous coronary intervention or coronary surgery. AMI patients requiring such care are typically transferred to the national cardiac center in Dar es Salaam.

### Participant Sampling and Recruitment

Purposive sampling was used to recruit participants across four key stakeholder groups: nurses, physicians, administrators, and patients from a variety of occupations, ages, genders, and educational backgrounds. Eligible participants were KCMC staff (physicians, nurses, administrators) who worked in the ED during the study period, as well as patients diagnosed with AMI in the KCMC ED during the study period. Individuals were excluded only if they were unable to provide informed consent.

Recruitment was conducted through direct invitations by study personnel, ensuring a diverse range of perspectives. Informed consent was obtained from all participants before data collection. Selected hospital staff were approached face-to-face by research staff during their break periods to offer participation, and selected patients were contacted via telephone to offer participation. All in-depth interview participants were compensated for their time (approximately 2 USD).

### Data Collection

Interviews were guided by a semi-structured interview guide, which was developed by the international, interdisciplinary investigator team (S2 text). These interview guides were informed by the Theoretical Framework for Acceptability (TFA), which encompasses seven domains of acceptability: affective attitude, burden, perceived effectiveness, ethicality, intervention coherence, opportunity costs, and self-efficacy.(15)The guide included open-ended questions exploring participants’ awareness and understanding of the intervention; perceived benefits and challenges; attitudes towards implementation and sustainability; and barriers and facilitators to intervention uptake. Interviews were conducted face-to-face in a private space in Swahili by Tanzanian research assistants with prior qualitative research experience who were not involved in participants’ clinical care. Interviewer training included qualitative interviewing techniques and study-specific protocol review. Each interview lasted approximately 45-60 minutes, and interviews continued until the study team determined thematic saturation had been reached. Repeat interviews were not conducted. All interviews were audio-recorded with participant consent, then transcribed and translated into English. Interviews were initially conducted with 20 participants (n=5 per stakeholder group). An interim thematic analysis was conducted by three members of the research team (JTH, TGT, SFS), who determined that thematic saturation had been achieved for each stakeholder group, so no additional interviews were conducted.

### Data Analysis

The qualitative data analysis for this study was conducted using an iterative, stepwise approach following the thematic framework described by *Applied Thematic Analysis*.(16) The study utilized an inductive approach to identify emergent themes related to intervention acceptability, informed by the TFA.(15) Interview transcripts were coded by trained research personnel with prior experience in qualitative analysis. Coders underwent additional training in thematic analysis principles prior to beginning formal analysis, and the coding team consisted of both cultural insiders (KFH, GLK, LW, JJM, PPS) and cultural outsiders (SS, JTH).

To ensure a rigorous and systematic approach, the coding process began with coders familiarizing themselves with a representative sample of interview transcripts. This initial phase allowed for an in-depth understanding of the interview format and content, ensuring alignment with the research objectives. Following this familiarization, a lead coder developed an initial codebook, informed by the domains of the TFA, that systematically categorized themes of acceptability into discrete codes. These codes identified the acceptability domain, whether a response indicated acceptability or unacceptability, and the specific reasoning underlying the respondent’s perspective. The codebook was iteratively refined throughout the coding process as emergent themes were identified through regular discussions between coders. All transcripts were independently coded by two members of the coding team, and coding discrepancies were reviewed and reconciled through group discussion and consensus.

Interviews were coded by respondent group to ensure that emerging themes could be captured within each stakeholder perspective. The codebook was expanded as necessary with each group, and modifications were made to reflect new themes. Each coder analyzed the interviews independently before convening to discuss discrepancies and reach a consensus. This process was repeated for all four stakeholder groups: nurses, administrators, physicians, and patients. The final codebook is provided in the Supporting Information (S3 File).

After a consensus was reached for all interviews, thematic analysis was conducted in NVIVO 14. High-level thematic analysis was led by two members of the research team (JTH and SS). Dominant themes were identified by respondent group and number of discrete respondents mentions. The final stage of analysis involved thematic synthesis, where patterns were identified across stakeholder groups to generate a comprehensive understanding of the intervention’s acceptability within the emergency department setting at KCMC.

### Reporting Standards

This study is reported in accordance with the Consolidated Criteria for Reporting Qualitative Research (COREQ) checklist (S1 Checklist).

### Ethical Considerations

Ethical approval was obtained from the Duke Health Institutional Review Board, the Kilimanjaro Christian Medical Centre Research Ethics Committee, and the Tanzania National Institutes for Medical Research Ethics Coordinating Committee. Written informed consent was obtained from all participants prior to their involvement. Confidentiality was maintained through de-identification of transcripts and secure data storage.

## Results

Twenty participants were enrolled in this study, including five physicians, five nurses, five administrators, and five patients (Table 1). The median age was 32 years (range 27–64), and 40% of participants were female. Additional participant characteristics are presented in Table 1.

**Table 1.**
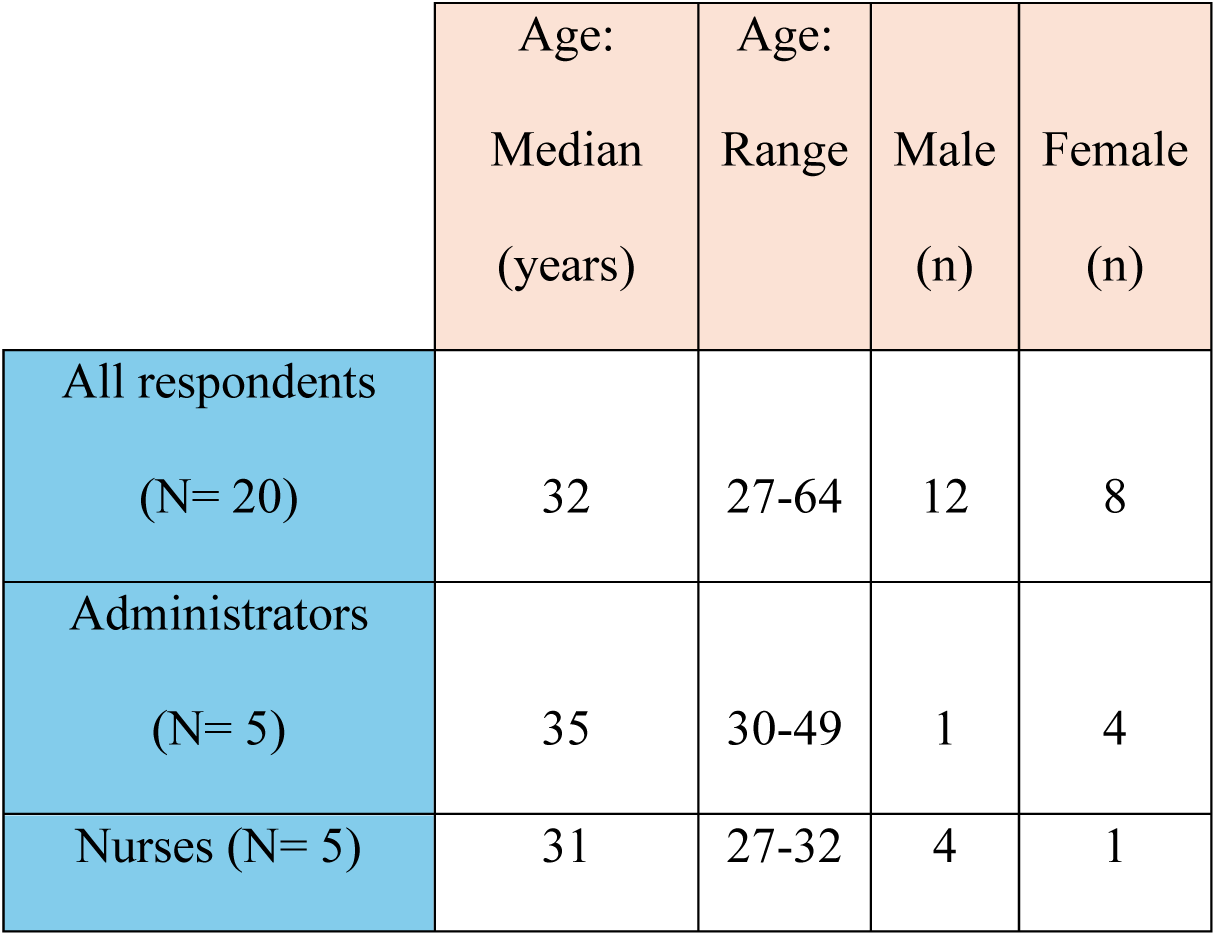
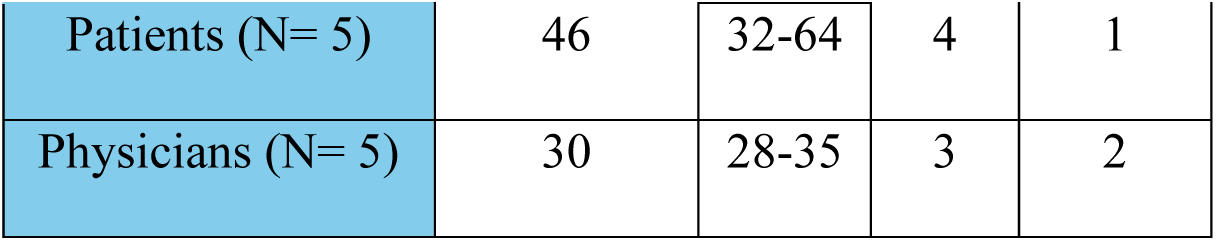
Participant characteristics, by stakeholder group (n = 20).

|  | Age:<br>Median<br>(years) | Age:<br>Range | Male<br>(n) | Female<br>(n) |
| --- | --- | --- | --- | --- |
| All respondents<br>(N= 20) | 32 | 27-64 | 12 | 8 |
| Administrators<br>(N= 5) | 35 | 30-49 | 1 | 4 |
| Nurses (N= 5) | 31 | 27-32 | 4 | 1 |
| Patients (N= 5) | 46 | 32-64 | 4 | 1 |
| Physicians (N= 5) | 30 | 28-35 | 3 | 2 |

Across stakeholder groups, we identified 19 dominant themes within all seven TFA domains (Table 2). Overall, participants reported positive affective attitudes toward the MIMIC intervention, describing it as minimally burdensome, well aligned with their professional and ethical values, and effective in improving AMI care. Staff highlighted increased awareness and confidence in diagnosing and treating AMI, while patients emphasized the value of the educational pamphlet in understanding and managing their condition. Divergent views emerged regarding the appropriateness of pamphlet distribution in the ED, representing the main area of disagreement across groups.

**Table 2.**
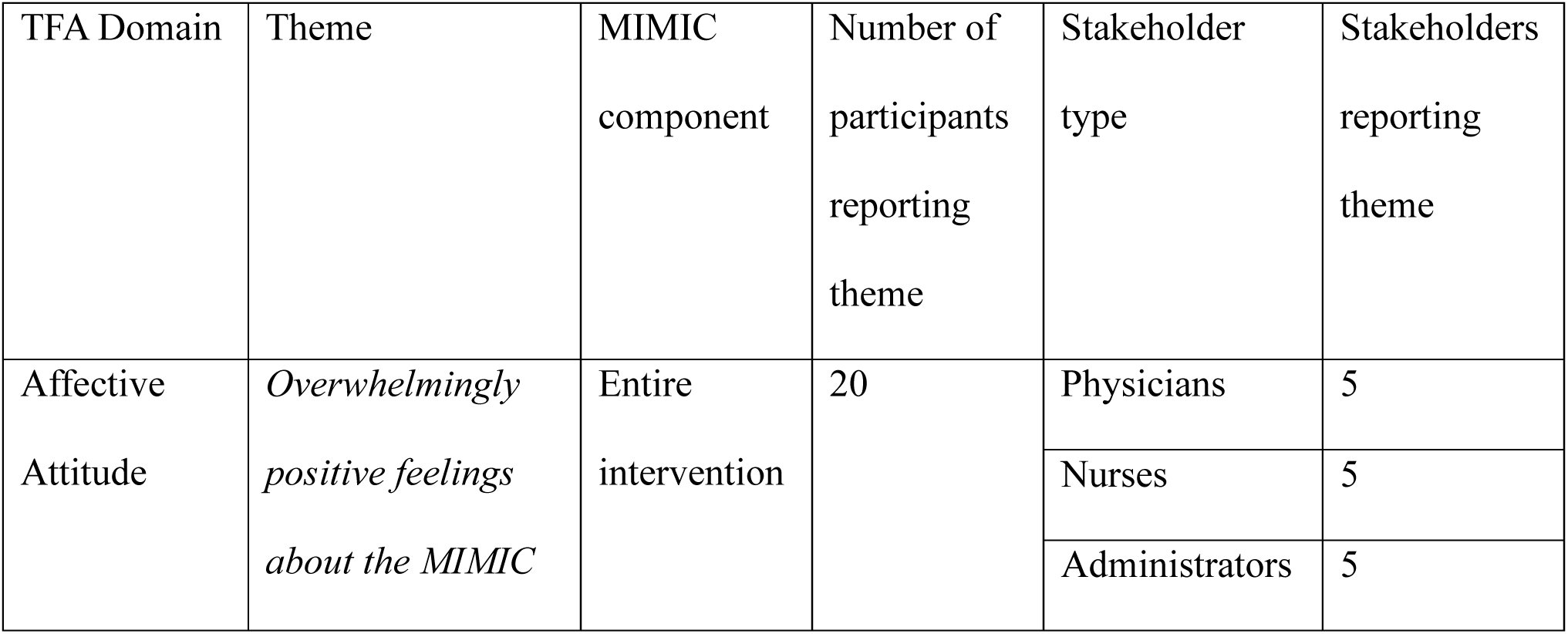
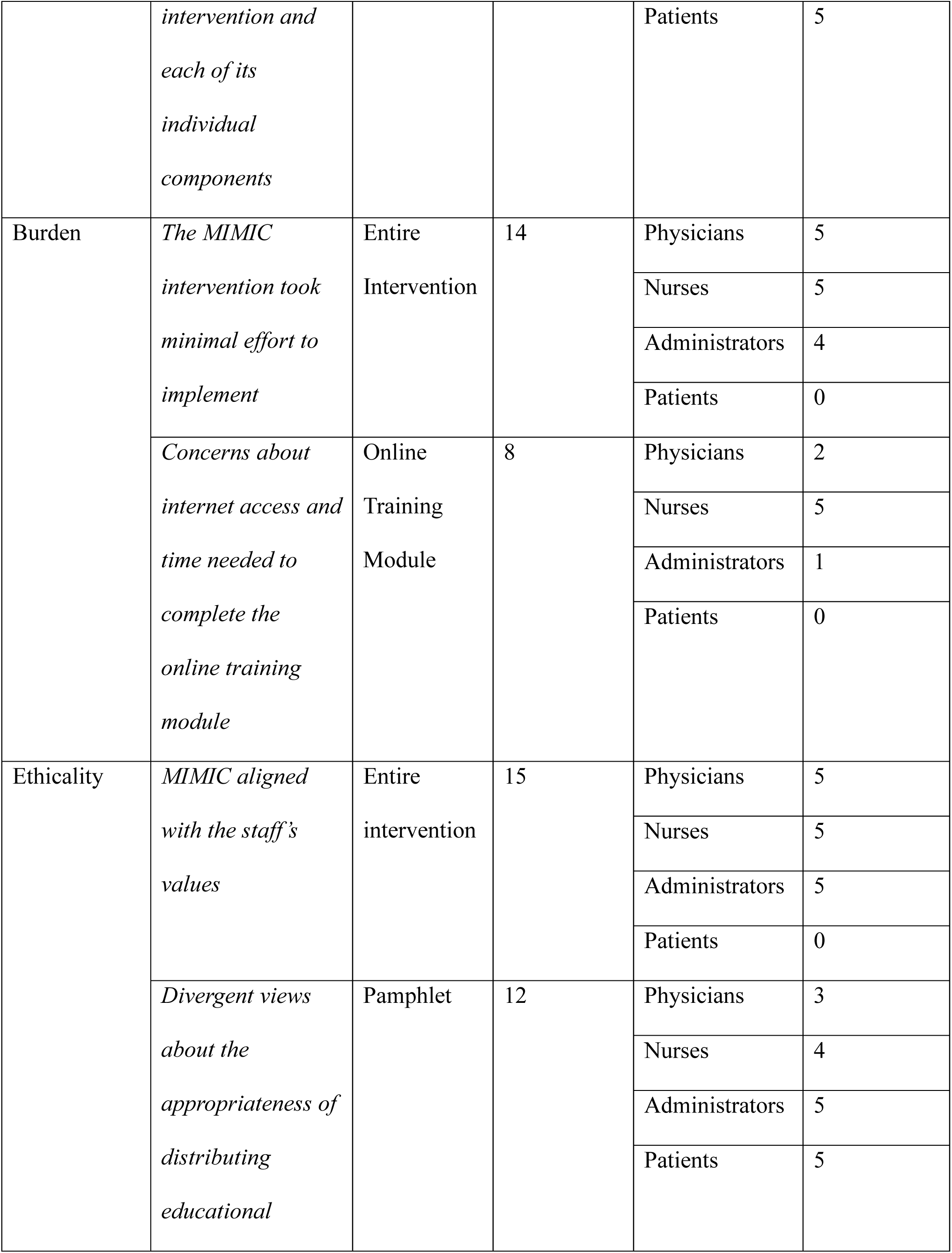
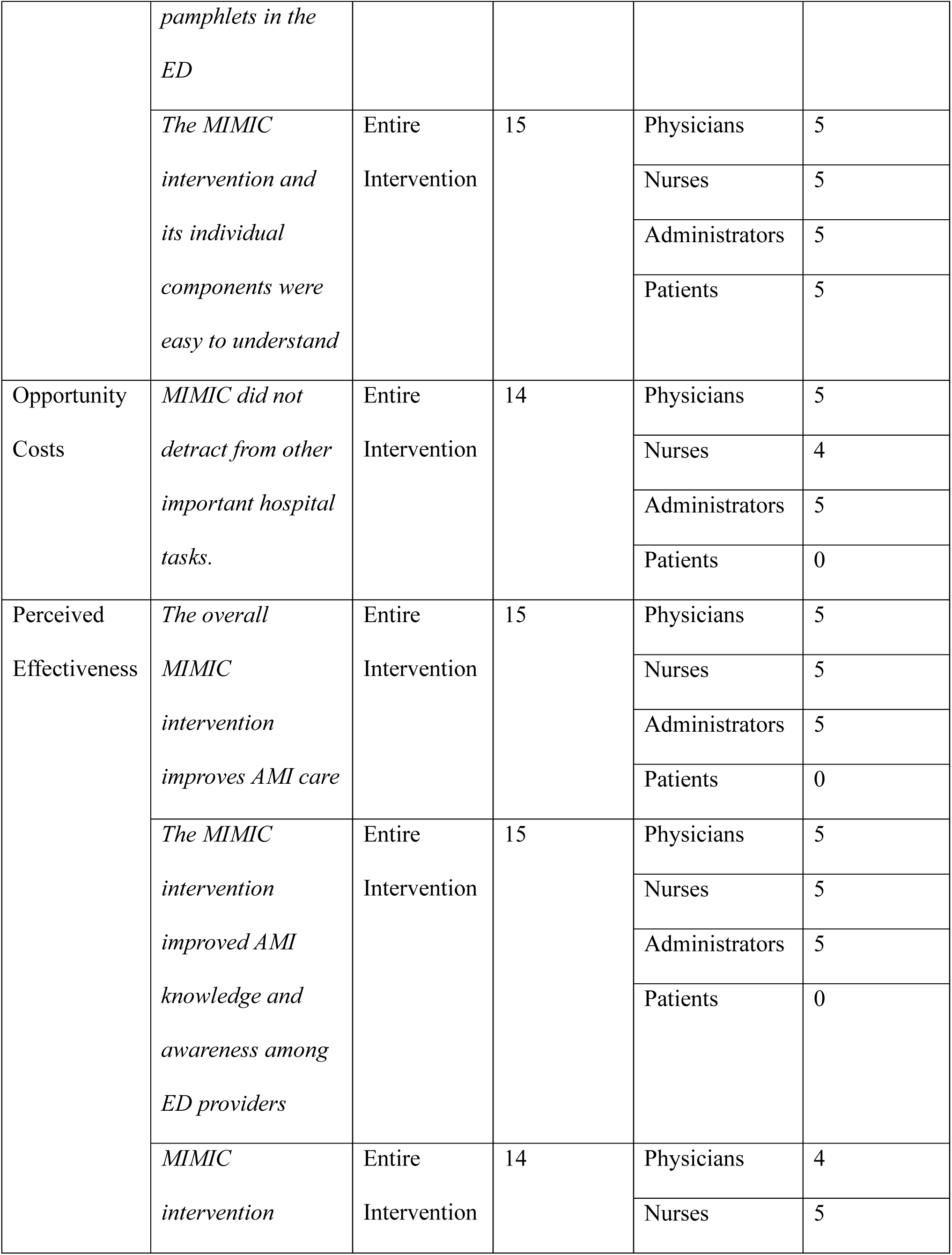
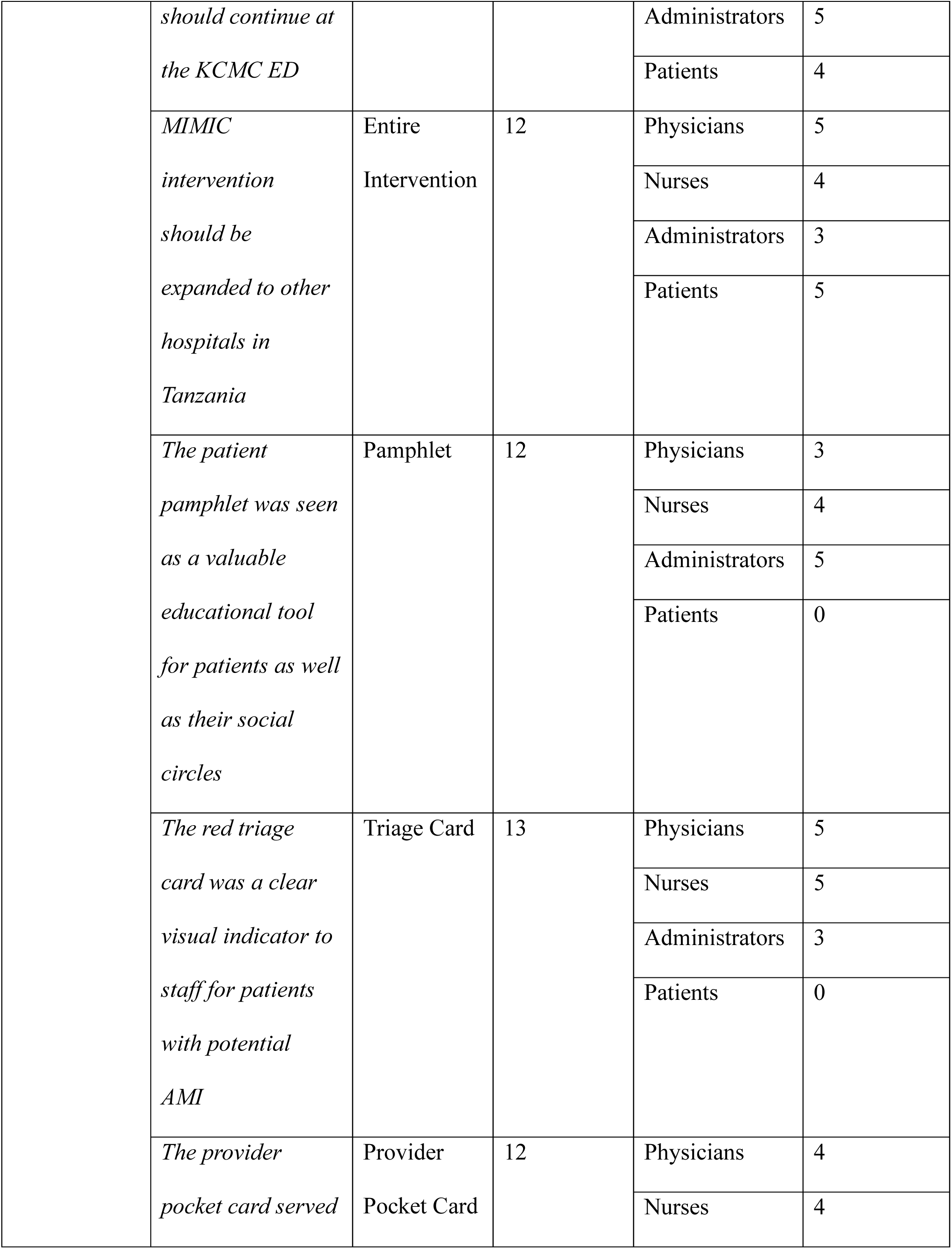
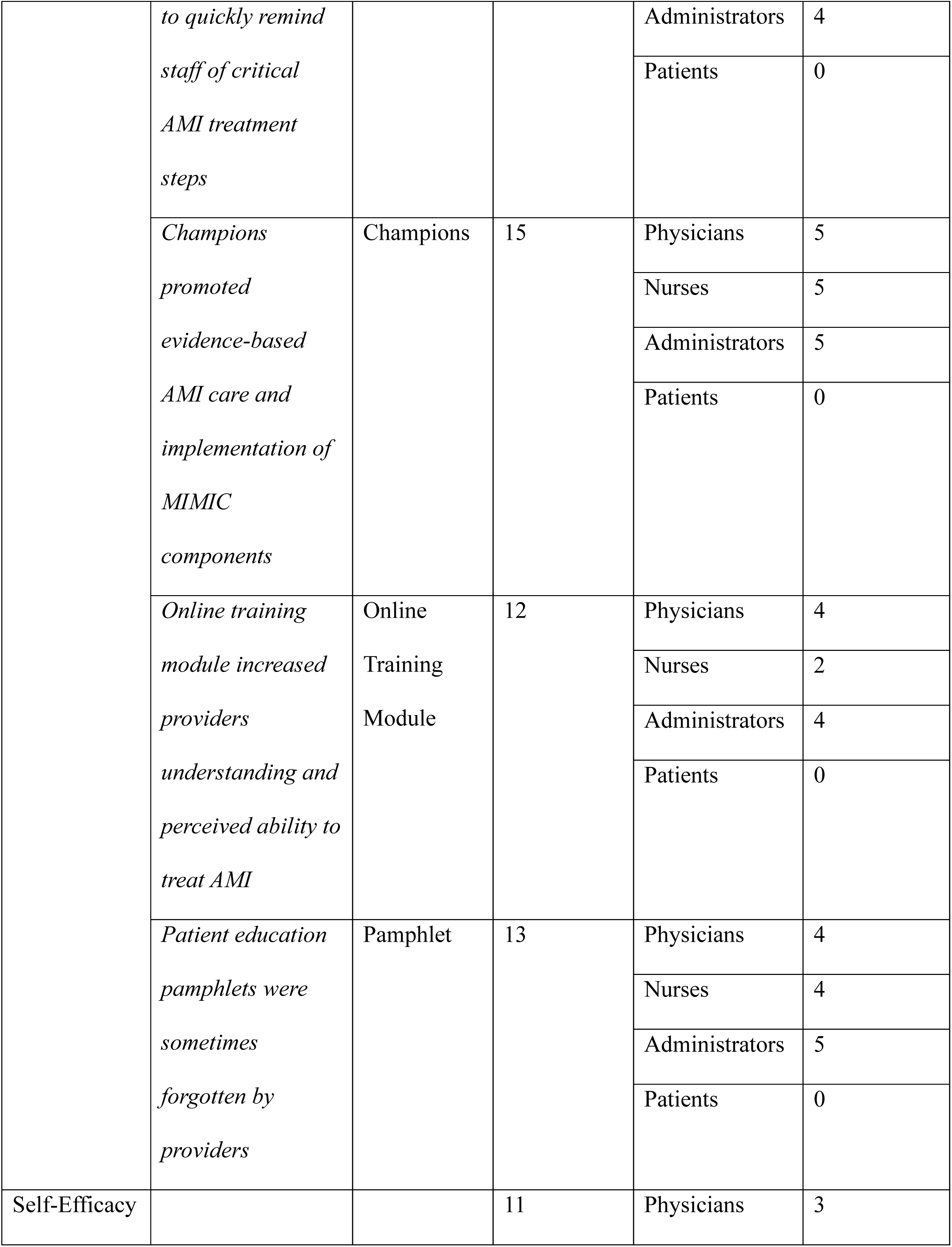
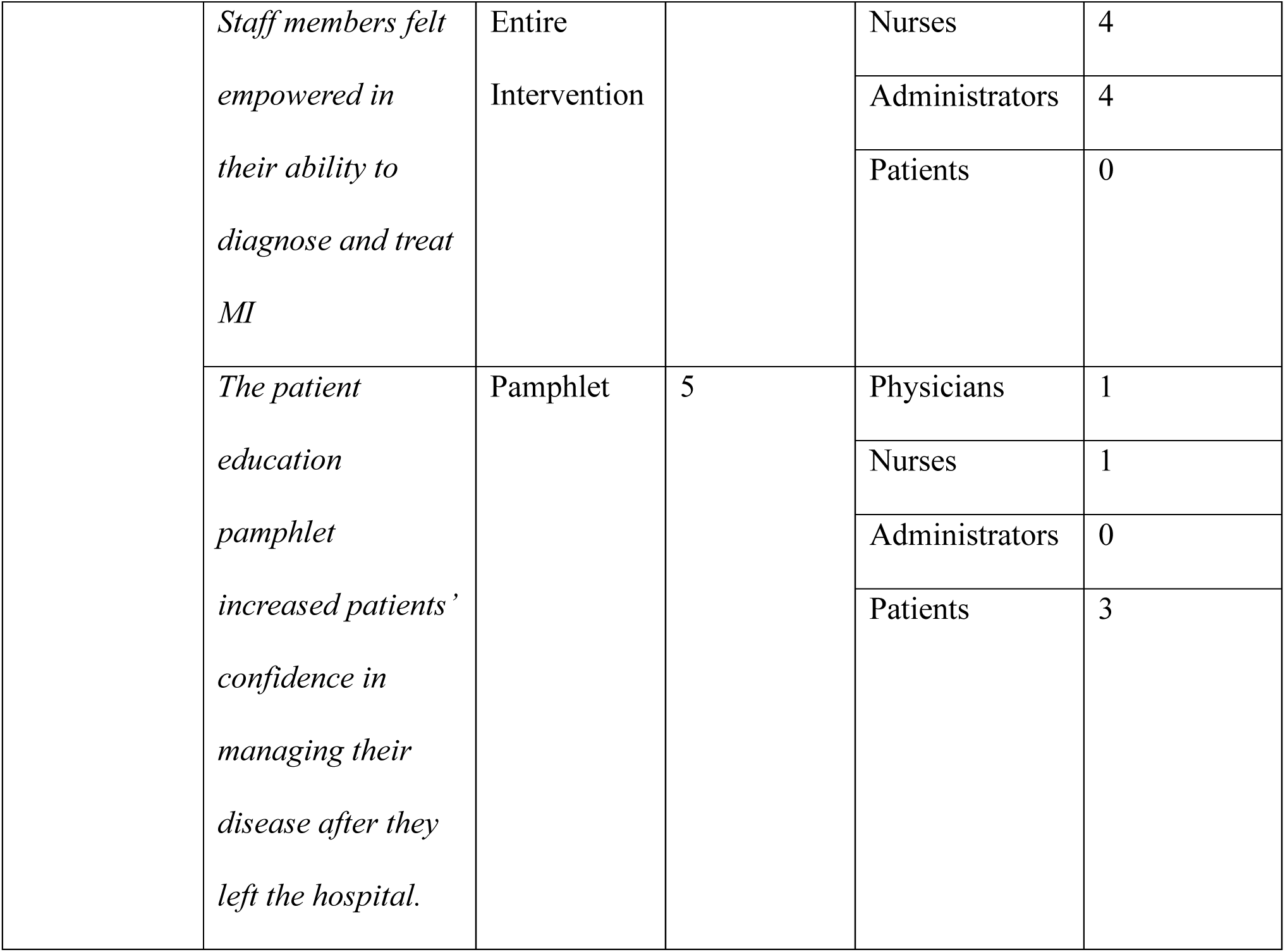
Dominant themes related to MIMIC acceptability, organized by Theoretical Framework of Acceptability domain, intervention component, and stakeholder group.

### Affective Attitude

#### Overwhelmingly positive feelings about the MIMIC intervention and each of its individual components

All participants across all stakeholder groups expressed positive affective attitudes towards the MIMIC intervention. Although some stakeholders had some reservations about certain components of the intervention, overall, all participants expressed positive sentiments towards MIMIC and its components. There was some variability in affective attitudes towards individual components across stakeholder groups. For example, as discussed below, patients expressed strong positive feelings about the educational pamphlets while feelings towards the pamphlets among administrators were more mixed. However, all respondents had a general overall positive affective attitude toward MIMIC, and no participant reported disliking MIMIC overall.

> “Yeah. I would frown filling in a death certificate but not MIMIC.”
>
> […]
>
> I have never seen a frown face when it’s come to MIMIC.” (Physician, M)”
>
> “I liked MIMIC. Not only liked it but I loved it! And I still love it!
>
> […] Everyone that I met thinks it is a very good intervention.” (Physician, F)

Patients, who only engaged with the educational pamphlet component of MIMIC, also reported uniformly positive feelings towards this intervention.

> I: “In general, how do you feel about this educational pamphlet?”
>
> R: “The pamphlet is good […]. They gave me that pamphlet, it helped me, and I received it because I knew that I already had MI and what was required for me to do, and at what time.” (Patient, M)

### Burden

#### The MIMIC intervention took minimal effort to implement

It was a widely held belief that MIMIC was not a burden to implement as an intervention because stakeholders saw improving quality of care to be part of their responsibilities as a healthcare provider. MIMIC was also noted to be a simple intervention with easily understandable components that do not require a large investment of time.

Moreover, providers felt that MIMIC, which was co-designed by KCMC ED staff and tailored specifically to the Tanzanian context, was designed to fit within existing workflows. For example, participants cited the way in which the “MI Suspect” red triage cards could be integrated seamlessly into the existing card-based triage system without adding burdensome additional tasks for staff. Furthermore, as the MIMIC intervention became normalized, participants reported that it became part of routine ED operations.

> I really don’t see the burden because it is not something completely new. We are using triage cards that we also had before. We have other cards that show if the patient has been seen, if they are awaiting responses and so on. Doing ECG and other tests are part of the doctor’s consultation. Doing troponin in the ED has also made things easier for us. Pocket cards do not give the doctors a burden but rather reminds them on what they are supposed to do. At the emergency we also have charts that remind us on different other diseases so it’s just similar. I don’t think they are a burden I think they are just part of normal routine. (Administrator, F)

> “MIMIC is not a hard thing to do. It is not burdensome. It doesn’t demand a lot from you, you just need to remember a few things. Do this, do that, remember the red tag and follow the protocol. Just a few things that don’t consume your time and energy.” (Physician, F)

#### Concerns about internet access and time needed to complete the online training module

Although participants overall felt that MIMIC was minimally burdensome, some respondents considered the online module somewhat burdensome to complete. A minor concern shared by nurses, administrators, and doctors was that an additional online training course was a burden because a busy clinical schedule necessitated the course to be completed on one’s own time outside of the hospital. There was no time allotted in regular working hours to complete the course. Completing the course at home also meant that a personal internet bundle was required, which brought up concerns about cost and internet access. Although free internet is available at the hospital and no respondent reported personally being unable to access the internet to complete the module, many participants expressed concerns about other staff members who might not have a smart phone or internet access.

> “This is dependent on the overlapping of tasks. You find that you have had a busy day at work, and you have been asked to do an online module. When you get home, you are exhausted, and all you want to do is just rest and say that you will do it on the next day. And that is the beginning of procrastination. Sometimes you cannot do it because of data and internet issues. When you ask some people to do it, they tell you that they do not have data and internet, or their phones are faulty. Those are some of the barriers” (Nurse, M)

### Ethicality

#### MIMIC aligned with the staff ‘s values

All groups interviewed believed that the MIMIC intervention aligned with their ethical values. Participants typically interpreted this alignment across two domains: hospital ethics and professional ethics. When discussing hospital values, respondents emphasized that as a tertiary care center, KCMC has a responsibility to recognize AMI and provide care that meets international standards. In terms of professional ethics, both nurses and physicians noted that MIMIC supported their core responsibility to save lives by enhancing their ability to do so.

> “I: Do you think the MIMIC intervention aligns with your values?
>
> R: Yes, it does completely because our aim in providing care is to get quality care. How is quality care described? Have we been able to follow the requirements to get quality care?
>
> So, I see it aligns with my values.” (Administrator, F)

#### Divergent views about the appropriateness of distributing educational pamphlets in the ED

Providers and patients had notably different perspectives on the appropriateness of distributing educational pamphlets to patients in the ED. Many ED staff members felt that the pamphlet should not be given to a patient while they are getting treated for a possible AMI as they are in distress and the additional information would be overwhelming and harmful to patients. Many providers stated that they thought the pamphlet should either be given later in the hospitalization or be distributed in community settings such as markets, churches, or mosques, rather than the ED. This sentiment was particularly common among departmental administrators. Beyond concerns about ED patients being too critically ill to receive the pamphlets, providers felt that ED doctors were too busy delivering life-saving care to these patients, resulting in the pamphlets often being forgotten or overlooked. These participants felt that doctors in less acute settings, such as the inpatient wards or outpatient clinics, would have more bandwidth to distribute the pamphlets.

> “You know in the case of emergencies, relatives or the patients are mostly in shock and unsettled. If you give them something to read, they keep it in their bags, they don’t read it. That’s my observation.” (Administrator, F)

> “As I mentioned earlier, most patients that come here are very old, very sick, in pain and uncooperative. So, we need to improve because we need to deal with the society that is caring for that patient and the population of society that is healthy, so that when they get that education, they may bring their patients early before they reach the end stage of AMI. Also, when the healthy society is educated, they will know how to prevent and control AMI. Not wait until when they reach that stage” (Administrator, F)

Notably, patients did not share any of these concerns. In addition to believing the pamphlets were effective (as described below), patients did not raise any ethical concerns about the timing or location of pamphlet distribution. Patients expressed uniformly positive perspectives on the ethicality of the pamphlets, stating that they helped them understand their disease and contributed to a sense of self-efficacy. Several patients referenced inadequate doctor-patient communication throughout their hospital stay and felt that the pamphlets addressed this information void.

> “[receiving the pamphlet in the hospital] matched my value because I received that pamphlet because I was diagnosed of a certain disease. After realizing that I had a certain a problem, the doctor explained it to me, I received the news and understood. Afterwards, he gave me that pamphlet which was like an addition to what he had already explained to me. I received the news verbally and in written form and it is better to have it in writing because you can keep that.” (Patient, M)

> “[Providing the pamphlet] is important. It is helpful, it gives people awareness regarding cardiovascular issues. I have been very sick and since I noted it as a problem, I have been hearing from people with MI that they are experiencing challenges like difficulty in breathing, or they have been hospitalized. It is very important and would be helpful if they continued providing them.” (Patient, M)

### Intervention Coherence

#### The MIMIC intervention and its individual components were easy to understand

Respondents stated that the MIMIC was easy to understand, and most could explain all components coherently without prompting. Respondents were aware of the both the stated goal of improving the diagnosis and treatment of AMI and how MIMIC accomplished this through its 5 components. As MIMIC was developed through a participatory co-design approach, several respondents noted being part of the development process. The written materials (patient pamphlet and provider pocket card) were noted to be clear and easily understood by both patients and providers due to their simple direct nature and clear Swahili.

> “MIMIC is an intervention that helps us identify and manage patients with myocardial infarction. Mostly identification of patients is done once they come into the ED with a red card, so we stay alert that this patient is a suspected case of MI. We also did an online course that helped us gain knowledge on myocardial infarction. We were also given pocket cards which reminded us of the management and diagnosis of myocardial infarction. Most of us have them in our pockets and phones. We also have patient education pamphlets that we give to the patient’s relatives which help shorten the explanation that you need to give to the patient or their relatives. After giving out those pamphlets, the doctor’s job is just to clarify on the things that they did not understand from the pamphlet. This will help increase patient’s knowledge on this disease. Champions also motivate us to ensure that we provide the appropriate management to MI patients and to remind the students and interns on the proper management of MI.
>
> […]
>
> “I don’t think that it’s a very difficult thing to understand” (Physician, F)

> “Honestly, the part that I understood really well is the [Patient education pamphlet] which discusses what MI is and how to prevent it for people without MI. How to avoid MI for example looking at the foods we eat, tobacco use and weight. It has really helped me as an MI patient. If someone asks me what causes MI, it’s easy for me to explain.” (Patient, M)

### Opportunity Costs

#### MIMIC did not detract from other important hospital tasks

Respondents stated that MIMIC did not take away from other important clinical tasks. The main reason cited was AMI is so dangerous and life-threatening that AMI care was the most important task to attend to. Thus, participants felt that MIMIC could not detract from the overall mission of the ED staff, which was to save their patients’ lives. Implementing MIMIC as a quality improvement project was also considered by staff members to be a core part of their job, so it was not considered to be in competition with other responsibilities. Moreover, since MIMIC required little additional time or effort in day-to-day care, participants did not feel it was likely to detract from other important responsibilities.

> “So, it can’t distract me from important work, there is nothing that is as important to me, as the patients during my shift. So as long as I am at the emergency department and patient comes in with acute coronary syndrome, I have to go through and adhere to MIMIC. It’s not a distraction, it just draws attention.” (Physician, F)

> “I: Have you ever thought that the MIMIC intervention may distract you from other important work?
>
> R: As I mentioned earlier, it is part of my work, so it doesn’t affect anything. Not only for me but also for my colleagues.” (Administrator, F)

### Perceived Effectiveness

#### The overall MIMIC intervention improves AMI care

Of all the domains of acceptability, perceived effectiveness was the most frequently mentioned and heavily emphasized element of MIMIC acceptability among participants. Doctors, nurses, and administrators strongly agreed that the MIMIC intervention substantially improved AMI care quality in the KCMC ED. Participants noted enhanced uptake of ECG and troponin testing, reducing missed AMI diagnoses, and increased use of evidence-based therapies like aspirin and clopidogrel, leading to improved patient survival. Acceptability of MIMIC was closely tied to its perceived effectiveness, which was frequently cited by participants as the primary reason for their approval. Staff expressed pride in these observable improvements, highlighting their improved ability to rapidly recognize AMI symptoms, use diagnostic tools, and make swift care decisions, such as transfers to specialized centers or thrombolytic administration. They also noted a significant cultural shift in the ED, with AMI now recognized as a critical emergency requiring immediate attention, contributing directly to better long-term survival outcomes.

> “Honestly, I see that we have gone a step ahead because some patients were mismanaged. People didn’t think it was important to give them Aspirin and Clopidogrel, let’s just give them one. But when you have [MIMIC], you can see. Is it important to do an ECG again? But [MIMIC] reminded us. I think now we are more able to identify the patients with this diagnosis compared to the previous years. […] So, it has given us a wider knowledge on how to manage and exclude other things that limited our management.” (Administrator, F)

> “I would say it effectively improved the care because yes, sometimes we miss [AMI cases] but now it’s very very few times. We recognize the cases super-fast and they get referred early and they get saved. Most of the cases.” (Physician, F)

> “It is good because it helps in identifying patients with AMI, which accounts for many lives. This problem was there and was hurting many people and many people died for that reason. Patients with AMI were coming to the emergency department and there were formerly many delays including delays in sending the patient to the ward. Now, we have reduced those delays and patients are rushed to [specialty cardiology hospital] with an ambulance and receive treatment on time. Of course, it helps because if MIMIC wasn’t there then we would be missing these diagnoses.” (Nurse, F)

### The MIMIC intervention improved AMI knowledge and awareness among ED providers

All administrators, nurses, and physicians felt that the MIMIC intervention improved AMI knowledge and awareness among all ED staff members. Participants felt that the intervention, particularly the online module and pocket cards, improved their personal knowledge of AMI which they felt led directly to improved AMI care. Participants thought MIMIC was particularly effective in dispelling common misconceptions, refreshing knowledge among experienced doctors, and educating new team members like interns.

> “I like MIMIC because it increases my awareness on this because initially, I did not know about AMI. It has explained AMI to me, and it still sharpens me. I can even educate community members on this now.” (Nurse, M)

> “For me, I am happy because it increases my knowledge. Let’s say I go and work in another hospital where such things have not arrived, I can act as a role model. If someone comes with a chest pain on the left side and it is radiating, I can tell them let’s start with this coming from the few things that I know.”
>
> (Nurse, F)

> “I: How about improvement of provider awareness? Do you think MIMIC has been effective at that?
>
> R: I’d personally say yes. If you compare the knowledge I had before this intervention and after, there’s a big gap.” (Physician, F)

#### MIMIC intervention should continue at the KCMC ED

Providers expressed a strong desire for the MIMIC intervention to continue in the KCMC ED after the study period completes. Staff members felt that MIMIC has allowed them to see how large of an issue AMI is in KCMC’s ED and thought it would lead to patient harm if the intervention were to be taken away. Many respondents also reported that they are actively maintaining the components of MIMIC and working to introduce them to new staff members as they arrive.

> “Please don’t stop this project, please don’t because we have only done it here at KCMC and we have realized that it is a leading cause of death. If you implement it in all hospitals in Tanzania, you will realize that many people die of myocardial Infarction.” (Administrator, F)

> “I: Do you think the MIMIC intervention can continue in the KCMC emergency department later on?
>
> R: Absolutely! We have seen that this is a good system. We are celebrating good behavior. We are maintaining it with both hands.” (Administrator, M)

> “I would like to thank you for this program because it has made a huge positive impact, something which we wish to continue. As I already mentioned, it shouldn’t end here but we should create an environment for those around us to understand this intervention so as to improve care for all patients.” (Nurse, M)

> “The major thing we are thinking about in the MIMIC intervention is for this project to continue because it reminds us on how to care for these patients. If you are a provider and have the ability and skills to care for these patients quickly, it will be a stable carrier full of success. Many of us want this to continue for many years.” (Physician, M)

> “It is important. Apart from personally liking the pamphlet, it is important to continue distributing it. It is helpful, it gives people awareness regarding cardiovascular issues. … It is very important and would be helpful if KCMC continued providing them.” (Patient, M)

#### MIMIC intervention should be expanded to other hospitals in Tanzania

In addition to stating that MIMIC should continue at KCMC, respondents expressed a desire to transplant the MIMIC intervention to other emergency departments across the country. Staff members felt that MIMIC should be spread not only to other large referral hospitals but also to smaller district hospitals and community clinics with less resources so that patients receive appropriate care without having to be referred to a larger hospital.

> “Many people will be helped. So many! Imagine people travelling from the villages to KCMC! How many hospitals have they skipped? How many hospitals have they been attended? They come here in the late stage. So MIMIC is something very nice, they should not stop. Please continue working on it.” (Administrator, F)

> “We are now aware of MI, and we need others to be aware. Tanzania is big, and we wish for patients to be diagnosed as soon as possible if they experience those symptoms. It is very important for this education to be taken to other emergency departments. Not only emergency rooms because some hospitals do not have an emergency department; those are the primary facilities that are attended by many patients who are poor. It should be provided to all facility levels and not only to the big hospitals.” (Physician, F)

> I am happy to be part of this intervention. If where we are planning to go [to other emergency departments] we will be received with open arms and then we will get more patients and we will reach more people.” (Administrator, M)

> “You should give pamphlets anywhere patients can be given them. Print out more copies and give the pamphlet to more people. This will help in reducing this problem. People will get to know the symptoms, the type and the numbers [of MI cases] will reduce.” (Patient, M)

#### The patient pamphlet was seen as a valuable educational tool for patients as well as their social circles

Patient participants felt that the educational pamphlets were very effective in educating not only themselves, but also their friends, neighbors, and relatives. Respondents reported that the pamphlets taught them about the symptoms and causes of AMI, a disease they had not heard of before. Many respondents stated that they used the pamphlet as a way to describe what AMI was to their loved ones, who may also have low health literacy, or as a form of community outreach to educate their social circles on the symptoms of AMI. The pamphlet was described in some instances as being the reason for bringing in a relative with symptoms of AMI to the hospital.

> “It has improved [things for me] because I didn’t know anything about that disease. When I was given the pamphlet, I read about the symptoms and causes and I understood that there is a disease of this kind.” (Patient, F)

> “The pamphlet is important. It is very important because the more you read it the more you remember. It reminds you the of symptoms, what to do, what to avoid to prevent another heart attack.” (Patient, M)

> “Honestly, I read this pamphlet with my children when it was given to me at KCMC. Everybody wanted to know what it was.” (Patient, M)

All the people that I have shared it with them were eager to read it. Some of them were eager to change because there are some things that they have seen there that cause MI. Some people have seen the need to reduce or stop for instance the smokers. I can see that some are trying to stop. Sometimes they fail but they are trying. The issue of obesity as well because I have spoken to many people and I see that they can understand. People like it, it is nice and some people have partners with this problem so they understand. (Patient, M)

### The red triage card was a clear visual indicator to staff for patients with potential AMI

The red card was noted to be useful as a clear visual indicator that prompted ED physicians to evaluate for AMI in their diagnostic workups. Staff appreciated the visually distinct nature of the card and commented that they could pick out the card clearly in a crowded ED. Staff also noted that they associated the algorithmic diagnostic workup of AMI with the red card and knew to check up on patients to see if the appropriate diagnostic tests have been administered or resulted. Physicians, nurses, and administrators all felt that AMI was commonly missed in the KCMC ED prior to MIMIC implementation primarily due to physicians failing to consider the diagnosis. Participants felt that the red triage card was particularly effective in addressing this gap because it made it nearly impossible for physicians not to consider AMI testing in patients with chest pain or dyspnea.

> “Basically, the card is unique, when someone sees it, it draws attention that there is something that needs to be done quickly or at a particular time.” (Nurse, M)

> “…the red card draws attention and even if you don’t explain it to providers, they already know that it is associated with MI. They are all focused on getting the ECG, Troponin, et cetera.” (Nurse, M)

> “The red card is a priority especially for providers working in the emergency department. That is like a visual language, before you say anything, there is already something in their minds to know that the particular patient might have MI.” (Nurse, M)

#### The provider pocket card served to quickly remind staff of critical AMI treatment steps

The provider pocket cards were lauded by the staff as a quick reminder for the information they learned previously during their training. Most providers described knowing the broad gist of AMI diagnosis and treatment but struggled with recalling the fine details, such as test cutoffs and medication doses. The pocket card served as an effective reminder and quick reference for those details. Although the pocket cards were provided both as physical laminated cards and digital copies, most providers preferred using the digital version on their phones. The pocket card was also noted to be of particular use to newer providers, including rotating interns, as a training tool on treatment algorithms and medication doses.

> “Pocket card is good because it is a small card with everything and makes things easier for us. It tells us what to do and what to prescribe. It helps even in reminding the new doctors that will come as well as the online training.” (Administrator, F)

> “The pocket cards, they really remind us. For me I usually confuse the doses for Aspirin and Clopidogrel. There is 300 or 600, so every time I will be peeping at the card and remember. Sometimes I forget to prescribe Heparin, I will peep here again and [chuckles] then I will remember.” (Physician, F)

#### Champions promoted evidence-based AMI care and implementation of MIMIC components

The Champions were seen both by all stakeholder groups as effective in promoting adherence to the MIMIC protocols and encouraging evidence-based AMI care overall. Several respondents stated that they were motivated to complete some of the more burdensome tasks, such as completing the online training, after being encouraged and reminded by the champions. Respondents noted that the value of the champions came from the constant reminders they provided during morning report and individual follow-ups about specific patient cases. Participants expressed appreciation for each responsibility of the champion role: ensuring staff are using the intervention components (such as triage cards, online module, educational pamphlets) appropriately, auditing care and providing follow-up to providers when care was sub-optimal, and publicly recognizing team members who delivered outstanding AMI care. Many providers thought the fact that the champions were already trusted colleagues and friends made them particularly effective in this role.

> ” They [the champions] are working well. We have different cases in the emergency department so sometimes it’s hard to remember all protocols, like doing an ECG and troponin in the first hour and again after three hours. It’s not easy if you don’t have someone who can remind you. You may find patients stay for more than three hours and are admitted with only a single ECG and troponin. So, it’s important for the champions to continue being there.” (Physician, M)

> “Because the champions were supervising the implementation of all the other components. So, without champions the rest probably couldn’t be implemented well. The champions were the ones following up on placing the cards, doing an ECG, reminding that when the troponin results come out to give Aspirin, do not be afraid of giving Aspirin to patients with CKD. Reminding us to do online module. I believe the champions were very important.” (Administrator, F)

#### Online training module increased providers understanding and perceived ability to treat AMI

Despite concerns about time and cost burdens, the online module was perceived by all groups to increase the ability of providers to diagnose and treat MI. Many respondents noted the specific direction on how to interpret tests that were previously not frequently administered such as troponins and ECG as well as specific direction on administering medication was the most beneficial. Participants also felt the module was effective addressing common misconceptions, such as concerns that aspirin was contraindicated in chronic kidney disease. The online module was also noted to be beneficial for providing feedback on what areas the respondent could improve on.

> “There are some medicines that we were so scared to prescribe but the online module assisted us in clearing those doubts.” (Nurse, F)

> “It (online module) updates us for instance, you usually submit your work, so they give us feedback on what we have done. What we have achieved, what needs improvement, and the like; so, it really helps us to know where we are, and we are going. We put effort on the points that we need to improve. It is really helpful.” (Nurse, F)

> “Because the online module has touched well on misconceptions, it ensures providers diagnose MI at an earlier stage. It also explains how to diagnose it from the ECG and troponin readings. It also explains what medicines to prescribe, at what dosage and at which time.” (Physician, M)

#### Patient education pamphlets were sometimes forgotten by providers

Respondents reported that patient education pamphlets were sometimes not given to patients, which limited their overall effectiveness. A common sentiment was that the busy workload of the emergency department distracted the staff from remembering to give the pamphlet to patients. ED staff also commented on the pamphlet’s storage location being out of sight and therefore difficult to remember. While several staff members commented on the fact that they thought the ED was the incorrect place to distribute the pamphlet (as discussed above), no person interviewed cited this as a reason they had not given a pamphlet to a patient.

> “There are two issues. You find that a person is too focused in providing care to give the pamphlet. The second thing is where they are stored. It becomes somewhat challenging to access them anytime. People don’t know where they are stored and by the time they know where the cards are the patient may have left. If it was kept in a place where everyone can see it would be easy to pick. At the emergency we have new doctors and nurses, we have new students, you understand. It becomes challenging to identify such things at that time. But if the cards are in an open space where everyone can see, When the student comes there, they will be asking what is this? They will be told that MI patients are given the pamphlet, so they know that if they get such patients, they need to give them.” (Nurse, M)

> “The first hassle is in looking for them because there are many files in there. We need to keep the educational pamphlets elsewhere please. You might look and miss them sometimes.” (Physician, F)

### Self-Efficacy

#### Staff members felt empowered in their ability to diagnose and treat MI

Respondents stated that the MIMIC intervention had boosted their confidence in their ability to manage patients with AMI. This confidence stemmed from a stronger knowledge base gained through training materials such as the online module and pocket card, which helped them recognize signs and symptoms and interpret diagnostic results. After MIMIC implementation, staff noted a significant improvement in their confidence to treat AMI, particularly in administering lifesaving medications. Prior to the intervention, guidelines were often not followed; however, afterward, participants expressed a clearer understanding of the importance of each medication. They also felt comfortable prescribing these treatments and teaching others to do the same.

> “I have proof, some patients I sent to [cardiac referral center] are still alive to this day. Most of my patients, all of them. I can say most of my patients they are doing good. I was able to diagnose them, give them the medication and send them for PCI and normal I make follow up to them. I did that.” (Physician, F)

> “I: After meeting the MIMIC training intervention, all the five parts, were you able to perform your part?
>
> R: Definitely! It has been like a wake-up call for everybody that MI is a serious thing. I can perform everything to ensure that everything is there, medication is there and investigations.” (Nurse, M)

> “Because one of the doctor’s values is being competent and objective. MIMIC intervention has been able to assist me with identifying these patients and channeling them to the appropriate places. It has increased something in my capacity to treat.” (Physician, M)

#### The patient education pamphlet increased patients’ confidence in managing their disease after they left the hospital

Patients stated that they had more confidence in their ability to manage their heart disease after they left the hospital due to the information provided in the patient education pamphlet. They repeatedly mentioned that the pamphlet clearly explained red flag symptoms and when to return to the hospital. They also emphasized that it described the medication regimen they should maintain after discharge, including when and how to take medications, and the timing of follow-up visits. Patients described the pamphlet as an excellent source of information and felt that it helped them manage their condition more effectively.

> “Because I want to know, what did they say on the pamphlet? When I felt the chest pain-like symptoms, I said what did they say here. I liked it because I know if I experience any problem, I will go back to the pamphlet to see what it says about that problem. For instance, if I get a headache I check to see if they mentioned headache as a symptom of heart attacks.” (Patient, M)

> “It has motivated me to continue using my medicines to cure this disease. If I use my medicines, it won’t be easy for that problem to recur.
>
> […]
>
> It has encouraged me to go to the clinic for follow up. That is why initially I was attending the monthly clinics, but I am now only having to go after every two months. I attend the clinics and do all tests. I do ECHO, ECG, every time that I go there I do all the tests. They check my progress and prescribe me medications, so it causes me to continue receiving care, like using the medicines, and preventing another heart attack. My health will become better, and I will be doing very fine.” (Patient, M)

## Discussion

Emergency medicine is an emerging field in SSA and Tanzania specifically.(17) In recent years there has increased effort to define and track quality metrics in EDs in SSA,(18) but few projects have progressed to implementation and subsequent evaluation. To our knowledge, MIMIC is the first quality improvement intervention for AMI care in SSA.(9) Using qualitative interviews from diverse stakeholders, this study builds on previous work to understand the acceptability of the MIMIC intervention among providers and patients and to generate actionable feedback to refine the intervention before planned expansion across Tanzania.

To evaluate the acceptability of this intervention, we used TFA, which has previously been used to evaluate acceptability of HIV-related healthcare interventions in SSA and in Tanzania specifically.(19–21) To our knowledge, this is the first use of TFA for ED-based implementation research in SSA; we found TFA to be a useful framework for obtaining thick description of intervention acceptability in our setting. Using the quantitative AIM instrument in provider and patients surveys, we previously reported that MIMIC was highly acceptable among patients and providers, with a mean AIM score of 4.82 out of 5 among providers and 4.68 out of 5 among patients.(11,12) However, such quantitative measures do not allow for the more nuanced understanding of intervention acceptability provided by TFA in the present study, such as divergent views on the appropriateness of the patient educational pamphlets or the strong link between perceived effectiveness and overall acceptability among providers. The limitations of AIM in understanding intervention acceptability have been widely reported,(11,22) and our study reaffirms the importance of qualitative methods in rigorous evaluation of implementation outcomes.

The MIMIC intervention was widely viewed as acceptable and effective by emergency department staff and patients at KCMC. Stakeholders reported positive attitudes across all constructs of the TFA, particularly perceived effectiveness. Perceived effectiveness was by far most emphasized domain of acceptability across participant interviews. Specific and detailed responses from all stakeholder groups pointed to meaningful improvements in AMI care as the basis for these views. Staff noted increased use of diagnostic tools such as ECG and troponin testing, improved recognition of AMI, and more consistent administration of aspirin and clopidogrel. These clinical gains were linked to specific intervention components such as the triage card, provider pocket card, and online training. Respondents described MIMIC as minimally burdensome due to its alignment with existing workflows. The red triage card, for example, fit seamlessly into the card-based triage system. While the online module required extra time and internet access outside working hours, it was still valued for improving provider knowledge. It is important to note that MIMIC was developed using a co-design process, in which Tanzanian ED physicians, nurses, and patients participated as equal partners in every step of intervention development and refinement.(8) This approach likely contributed to the broad acceptability of MIMIC among providers and patients, as the input of intended users in the design process likely promoted usability and minimized burden.(23,24)

Ethical alignment was another key driver of acceptability as participants viewed MIMIC as consistent with their duty to provide high-quality care. However, there were differing views on the appropriateness of distributing patient pamphlets in the ED. Staff worried that acutely ill patients could be overwhelmed, while patients consistently found the pamphlets empowering and informative. Staff concerns about the educational pamphlet likely contributed to limited fidelity to this component of the MIMIC intervention; in the pilot trial, only 38% of patients with AMI in the KCMC ED actually received the pamphlet.(10) This study identified an important need to modify the process for distributing the patient educational pamphlets, which were often forgotten by ED providers. Possible solutions include identifying a designated ED staff person (likely a non-physician) to distribute these pamphlets or shifting this task responsibility to the inpatient team.

Patients were only able to comment on the patient education pamphlet, as it was the only component they interfaced with, but they overwhelmingly expressed positive sentiments about its educational value. This stood in contrast to provider concerns that patients might not read the pamphlet. However, no patient interviewed reported not reading pamphlet. Notably, we only interviewed patients who had received the pamphlets since other patients would not be able to comment on the acceptability of the pamphlets. This approach may have biased our results, as only a minority of patients with AMI actually received the pamphlets.(10) If providers were less likely to give pamphlets to patients who would have been less receptive to the pamphlets, then the patient perspectives elicited in this study may not be representative of the acceptability of the pamphlets in the general patient population. Additional study is needed to explore acceptability of the pamphlets in a broader patient population, including among patients who have limited literacy.

This study had several limitations. One limitation was its single-site design in a hospital closely involved with MIMIC’s development, which may have introduced bias, limiting the generalizability of our findings. Future studies should investigate acceptability of the MIMIC intervention in diverse EDs across Tanzania. Additionally, like all qualitative studies, participant responses may have been subject to social desirability bias, which could skew responses toward more positive affective expressions.To mitigate this potential bias, interviewers underwent extensive training in best practices in in-depth interview techniques to create a non-judgmental environment and encourage honest, critical feedback. Nonetheless, the consistency and specificity of responses, support the credibility of the findings.

In conclusion, in an ED in northern Tanzania, the MIMIC intervention was highly acceptable across all TFA domains among providers and patients. Future research should evaluate intervention acceptability, feasibility, and effectiveness in EDs across Tanzania.

## Supporting information

S1 Checklist

## Data Availability

The qualitative data supporting the findings of this study contain potentially identifiable human participant information and cannot be made publicly available. De-identified excerpts relevant to the study findings are included in the manuscript. Additional data may be made available upon reasonable request to the Duke University Institutional Review Board and the Kilimanjaro Christian Medical Centre Research Ethics Committee for researchers who meet criteria for access to confidential data.

## Acknowledgements

We gratefully acknowledge the KCMC ED staff for their participation and collaboration in this study. We gratefully acknowledge Godfrey Kweka, Jerome Mlangi, Tumsifu Tarimo, Pankrasi Shayo and Kelvin Haukila for serving as research assistants and collecting the data for this study.

## Supporting Information

**S1 Checklist. COREQ checklist.** Consolidated Criteria for Reporting Qualitative Research (COREQ) checklist for this study.

**S2 Text. Semi-structured interview guides.** Semi-structured interview guides used for in-depth interviews with patients, providers, and administrators participating in the MIMIC intervention, including questions assessing intervention acceptability and feasibility.

**S3 File. Qualitative codebook.** Final codebook used to code transcripts and organize themes mapped to the Theoretical Framework of Acceptability.

## References

1. Global Health Estimates 2021: Deaths by Cause, Age, Sex, by Country and by Region, 2000-2021. [Internet]. Geneva, World Health Organization; 2024. Report No. Available from: https://www.who.int/data/gho/data/themes/mortality-and-global-health-estimates/ghe-leading-causes-of-death#:∼:text=The%20world’s%20biggest%20killer%20is,9.0%20million%20deaths%20in%202021

2. Hertz JT, Madut DB, Rubach MP, William G, Crump JA, Galson SW, et al. Incidence of Acute Myocardial Infarction in Northern Tanzania: A Modeling Approach Within a Prospective Observational Study. J Am Heart Assoc. 2021 Aug 3;10(15):e021004. doi:10.1161/JAHA.121.021004 PubMed PMID: 34320841; PubMed Central PMCID: PMC8475708.

3. Yao H, Ekou A, Niamkey T, Hounhoui Gan S, Kouamé I, Afassinou Y, et al. Acute Coronary Syndromes in Sub-Saharan Africa: A 10-Year Systematic Review. JAHA. 2022 Jan 4;11(1):e021107. doi:10.1161/JAHA.120.021107

4. Hertz JT, Sakita FM, Prattipati S, Coaxum L, Tarimo TG, Kweka GL, et al. Improving acute myocardial infarction care in northern Tanzania: barrier identification and implementation strategy mapping. BMC Health Serv Res. 2024 Mar 28;24(1):393. doi:10.1186/s12913-024-10831-5

5. Hertz JT, Kweka GL, Manavalan P, Watt MH, Sakita FM. Provider-perceived barriers to diagnosis and treatment of acute coronary syndrome in Tanzania: a qualitative study. Int Health. 2020 Feb 12;12(2):148–54. doi:10.1093/inthealth/ihz061 PubMed PMID: 31329876; PubMed Central PMCID: PMC7017879.

6. Goli S, Sakita FM, Kweka GL, Tarimo TG, Temu G, Thielman NM, et al. Thirty-day outcomes and predictors of mortality following acute myocardial infarction in northern Tanzania: A prospective observational cohort study. International Journal of Cardiology. 2021 Nov;342:23–8. doi:10.1016/j.ijcard.2021.08.002

7. Dennis JA, Zhang Y, Zhang F, Kopel J, Abohelwa M, Nugent K. Comparison of 30-day mortality and readmission frequency in women versus men with acute myocardial infarction. Baylor University Medical Center Proceedings. 2021 Nov 2;34(6):668–72. doi:10.1080/08998280.2021.1945364

8. Hertz JT, Stark K, Sakita FM, Mlangi JJ, Kweka GL, Prattipati S, et al. Adapting an Intervention to Improve Acute Myocardial Infarction Care in Tanzania: Co-Design of the MIMIC Intervention. Ann Glob Health. 2024;90(1):21. doi:10.5334/aogh.4361 PubMed PMID: 38495415; PubMed Central PMCID: PMC10941691.

9. Hertz JT, Sakita FM, Rahim FO, Mmbaga BT, Shayo F, Kaboigora V, et al. Multicomponent Intervention to Improve Acute Myocardial Infarction Care in Tanzania: Protocol for a Pilot Implementation Trial. JMIR Res Protoc. 2024 Sep 24;13:e59917. doi:10.2196/59917

10. Hertz JT, Sakita FM, Munshi ZR, Rahim FO, Mganga D, Kachenje A, et al. Implementation Outcomes of an Intervention to Improve Myocardial Infarction Care in Tanzania. Ann Glob Health. 2025;91(1):43. doi:10.5334/aogh.4651 PubMed PMID: 40778095; PubMed Central PMCID: PMC12330802.

11. Weiner BJ, Lewis CC, Stanick C, Powell BJ, Dorsey CN, Clary AS, et al. Psychometric assessment of three newly developed implementation outcome measures. Implementation Sci. 2017 Dec;12(1):108. doi:10.1186/s13012-017-0635-3

12. Hertz JT, Sakita FM, Haukila KF, Shayo PS, Shayo FM, Willy J, et al. Acceptability and feasibility of a multicomponent intervention to improve acute myocardial infarction care in Northern Tanzania: The MIMIC pilot trial. PLoS One. 2025;20(9):e0333271. doi:10.1371/journal.pone.0333271 PubMed PMID: 40996964; PubMed Central PMCID: PMC12463254.

13. Wang C, Sakita FM, Sumner S, Shayo FM, Martin Z, Msangi W, et al. Sustainability and normalization of an intervention to improve evidence-based myocardial infarction care in Tanzania. Implement Sci Commun. 2026 Jan 15;7(1):27. doi:10.1186/s43058-026-00860-y

14. Hertz JT, Nworie JE, Shayo F, Galson SW, Coaxum LA, Daniel I, et al. Acute myocardial infarction diagnosis and treatment following implementation of a multicomponent intervention in Tanzania: the MIMIC pilot trial. BMJ Open. 2025 Nov;15(11):e107857. doi:10.1136/bmjopen-2025-107857

15. Sekhon M, Cartwright M, Francis JJ. Acceptability of healthcare interventions: an overview of reviews and development of a theoretical framework. BMC Health Serv Res. 2017 Dec;17(1):88. doi:10.1186/s12913-017-2031-8

16. Guest G, MacQueen K, Namey E. Applied Thematic Analysis [Internet]. 2455 Teller Road, Thousand Oaks California 91320 United States: SAGE Publications, Inc.; 2012 [cited 2026 Feb 28]. Available from: https://methods.sagepub.com/book/applied-thematic-analysis doi:10.4135/9781483384436

17. Mould-Millman NK, Dixon J, Burkholder TW, Sefa N, Patel H, Yaffee AQ, et al. Fifteen years of emergency medicine literature in Africa: A scoping review. African Journal of Emergency Medicine. 2019 Mar;9(1):45–52. doi:10.1016/j.afjem.2019.01.006

18. Broccoli MC, Moresky R, Dixon J, Muya I, Taubman C, Wallis LA, et al. Defining quality indicators for emergency care delivery: findings of an expert consensus process by emergency care practitioners in Africa. BMJ Glob Health. 2018 Feb;3(1):e000479. doi:10.1136/bmjgh-2017-000479

19. Sekhon M, Van Der Straten A, on behalf of the MTN-041/MAMMA Study Team. Pregnant and breastfeeding women’s prospective acceptability of two biomedical HIV prevention approaches in Sub Saharan Africa: A multisite qualitative analysis using the Theoretical Framework of Acceptability. Dubé K, editor. PLoS ONE. 2021 Nov 16;16(11):e0259779. doi:10.1371/journal.pone.0259779

20. Pamba D, Sanga E, Mlalama K, Maganga L, Mangu C, Lwilla A, et al. Optimizing delivery strategies for 3HP TB preventive treatment in Tanzania: A qualitative study on acceptability of family approach in HIV care and treatment centers. Robinson J, editor. PLOS Glob Public Health. 2024 Dec 17;4(12):e0003142. doi:10.1371/journal.pgph.0003142

21. Shayo EH, Kivuyo S, Seeley J, Bukenya D, Karoli P, Mfinanga SG, et al. The acceptability of integrated healthcare services for HIV and non-communicable diseases: experiences from patients and healthcare workers in Tanzania. BMC Health Serv Res. 2022 Dec;22(1):655. doi:10.1186/s12913-022-08065-4

22. Hamm RF, Levine LD, Szymczak JE, Parry S, Srinivas SK, Beidas RS. An innovative sequential mixed-methods approach to evaluating clinician acceptability during implementation of a standardized labor induction protocol. BMC Med Res Methodol. 2023 Aug 29;23(1):195. doi:10.1186/s12874-023-02010-7

23. Slattery P, Saeri AK, Bragge P. Research co-design in health: a rapid overview of reviews. Health Res Policy Sys. 2020 Dec;18(1):17. doi:10.1186/s12961-020-0528-9

24. Silvola S, Restelli U, Bonfanti M, Croce D. Co-Design as Enabling Factor for Patient-Centred Healthcare: A Bibliometric Literature Review. CEOR. 2023 May;Volume 15:333–47. doi:10.2147/CEOR.S403243

