## Supplementary material for "Acceptability of an intervention to improve uptake of evidence-based emergency myocardial infarction care in Tanzania: A qualitative study": S2 Text

**MIMIC Acceptability & Feasibility In-Depth Interviews**

**INTERVIEW GUIDE FOR PROVIDERS & ADMINISTRATORS**

Introduction: As you know, the KCMC EMD Team has worked to create a quality improvement intervention to improve MI care, called “MIMIC”. This intervention consisted of the following: (1) use of special red “Emergency ACS” cards by the triage nurses to indicate patients with possible ACS, (2) an online module with ACS refresher training for all EMD providers, (3) pocketcards for EMD doctors to use to help them remember ACS diagnosis and treatment, (4) educational pamphlets for patients with ACS, and (5) a designated physician champion and nurse champion to help encourage the team to improve ACS care.

You are being asked to participate in this study because you are a provider or have an administrative role within the KCMC ED. We are asking for your perspective on the MIMIC Intervention.

Background Information

1. Can you please share your age, gender, and role within KCMC EMD?
2. Can you please share your training background and years of practice?
3. How long have you been working in the KCMC ED?

Let’s start by talking about the MIMIC intervention.

4. Can you explain the intervention to me in your own words?
   a. What parts of the intervention do you think you understood the best? Why?
   b. Which parts of the intervention did you understand the least? Why?
   c. After getting trained in the MIMIC intervention, were you able to perform your parts of the intervention (triage cards, pocket cards, educational pamphlets, online module, champion) by yourself?
   d. If you needed help, what help did you need?
   e. How much training did you need to learn how to do MIMIC? How much time did this training take place?
5. Overall how do you feel about the MIMIC Intervention?
   a. Why do you feel that way?
   b. How do you feel about the triage red card system? Why?
   c. How do you feel about the pocket cards? Why?
   d. How do you feel about the patient educational pamphlets? Why?
   e. How do you feel about the champions? Why?
   f. How do you feel about the online training module? Why?
   g. Overall, did you like the MIMIC intervention? Why?
   h. Which part of the MIMIC intervention was your favorite? Why?
   i. Which part of the MIMIC intervention was your least favorite? Why?
   j. Would you be happy to see the MIMIC intervention continue in the ED?
   k. What do you think other people think about the MIMIC intervention?
   l. Why do you think that?
6. Do you think the MIMIC intervention created any burdens for KCMC staff?
   a. Why do you think that?
   b. What were the most burdensome part(s) of the intervention? Why those parts?
   c. What were the least burdensome part(s) of the intervention? Why?
   d. Did the MIMIC intervention consume too much time for staff? Why?
   e. Did the MIMC intervention create any hassles for staff? Tell me more.
7. Do you think the MIMIC intervention had other negative consequences by drawing time, attention, or resources from other important priorities?
   a. Why?
   b. What other priorities, projects, tasks do you think were negatively impacted by the MIMIC intervention? Why?
   c. Did you ever feel like the MIMIC intervention distracted you from other important work? Tell me more about that.
   d. Overall, do you think the MIMIC intervention was worth it or do you think it caused more harm than good? Why do you say that?

Acceptability: Ethicality

8. Do you think the MIMIC intervention was ethical?
   a. Why or why not?
   b. Do you think the MIMIC intervention aligned with your values? Why?
   c. Do you think the MIMIC intervention aligned with the hospital’s overall values? Why or why not?
   d. Were there any specific parts of the MIMIC intervention that aligned well with your values? Why?
   e. Were there any specific parts of the MIMIC intervention that didn’t align well with your values? Why?

Let’s talk whether you feel the intervention is working.

9. Do you think the MIMIC intervention was effective in improving MI care?
   a. Why?
   b. Do you think the MIMIC intervention improved diagnosis of MI here? Why?
   c. Do you think the MIMIC intervention improved treatment of MI here? Why?
   d. Do you think the MIMIC intervention improved provider awareness? Why?
   e. Do you think the MIMIC intervention improved patient education? Why?
   f. We talked about five different parts of the MIMIC intervention. What were the most effective parts of the MIMIC intervention? Why?
   g. What were the least effective parts of the MIMIC intervention? Why?
   h. What could be changed about the MIMIC intervention to make it more effective?
10. Do you think you were able to participate in the MIMIC intervention successfully?
    a. Why?
    b. We have the online training module, pocket cards patient educational pamphlets and champions (and for nurses add red triage cards). Did you encounter any challenges?
    c. Anything else?

Feasibility

11. Do you think the MIMIC intervention overall was feasible?
    a. Why?
    b. Do you think the KCMC staff can continue to do the MIMIC intervention in the future? Why?
    c. What parts of MIMIC make this most likely to be used at KCMC even after the study is finished?
    d. When the study is finished, which parts of MIMIC do you think KCMC staff might find hard and start to do less often?
    e. Do you think the MIMIC intervention could work in other emergency departments in Tanzania? Why or why not?
    f. What would have to change about MIMIC so that other emergency departments could use it easily and with every eligible patient?

Conclusion

12. Thank you for your time. Do you have any other thoughts you would like to share with me before we conclude?

**MIMIC Acceptability & Feasibility In-Depth Interviews**

**INTERVIEW GUIDE FOR PATIENTS**

Introduction: The KCMC EMD Team has worked to create a quality improvement intervention to improve MI care, called “MIMIC”. This intervention consisted of multiple components, including educational pamphlets for patients with heart attacks.

You are being asked to participate in this study because you recently received an educational pamphlet about heart attacks from the KCMC ED

Background Information

13. Can you please share your age and gender, and role?
14. Can you please share your educational background and current occupation?
15. Where do you currently live?

Engagement

16. Did you read the pamphlet? Why or why not?

Acceptability: Self-efficacy

17. Do you think you were able to read and understand the pamphlets successfully?
    a. Why?
    b. Did you encounter any challenges?

Acceptability: Ethicality

18. Do you think the pamphlets aligned with your values?
    a. Why or why not?

Acceptability: Intervention Coherence

19. Do you think you understood the pamphlets?
    a. Why?
    b. Can you explain what the pamphlet said in your own words?
    c. What parts of the pamphlet do you think you understood the best? Why?
    d. Which parts of the pamphlet did you understand the least? Why?
    e. What questions do you have about the pamphlets?

Acceptability: Perceived effectiveness

20. Do you think the pamphlets were effective in educating you about heart attacks?
    a. Why?
    b. Do you think the pamphlets improved your understanding of what a heart attack is? Why?
    c. Do you think the pamphlets improved your understanding of the treatment of heart attacks? Why?
    d. Do you think the pamphlets will make it more likely that you take your medications as prescribed? Why?
    e. Do you think the pamphlets will make it more likely that you attend your follow-up appointments as instructed? Why?
    f. What were the most effective parts of the pamphlets? Why?
    g. What were the least effective parts of the pamphlets? Why?
    h. What could be changed about the pamphlet to make it more effective?

Acceptability: Affective Attitude

21. Overall how do you feel about the educational pamphlet?
    a. Why do you feel that way?
    b. What do you think other patients would think about the pamphlet?
    c. Why do you think that?
    d. Overall, did you like the pamphlets intervention? Why?
    e. Would you be happy to see the pamphlets continue to be given to patients in the KCMC ED?

Acceptability: Burden

22. Do you think the pamphlets created any burdens for you or your family?
    a. Why do you think that?
    b. Did the pamphlets create any hassles for you or others? Why?

Acceptability: Opportunity costs

23. Do you think the pamphlets had negative consequences by drawing time, attention, or resources from other important priorities?
    a. Why?
    b. Overall, do you think the pamphlets were worth it or do you think it caused more harm than good? Why do you say that?

Feasibility

24. Do you think the educational pamphlets were feasible?
    a. Why?
    b. What would you change about pamphlets to make it more feasible?
    c. How much time did you spend reading the pamphlets?
    d. Do you think the KCMC staff can continue distributing pamphlets in the future? Why?
    e. Do you think the pamphlets could work in other emergency departments in Tanzania? Why or why not?

Conclusion

25. Thank you for your time. Do you have any other thoughts you would like to share with me before we conclude?
